# Association of Objective Long Sleep Duration and Insomnia with Objective Short Sleep Duration Phenotypes with Mortality in Older Persons

**DOI:** 10.1101/2025.10.27.25338896

**Authors:** Brienne Miner, Yulu Pan, Gawon Cho, Melissa Knauert, Henry K. Yaggi, Katie Stone, Jamie M. Zeitzer, Kristine Ensrud, Sonia Ancoli-Israel, Susan Redline, Kristine Yaffe, Margaret Doyle

## Abstract

**Objective:** To investigate the association of objective long sleep duration (LS) and insomnia with objective short sleep duration (ISSD) with mortality in older persons.

**Methods:** In 3,054 men (average age 76.4±5.5; mean follow-up=12.1 years) and 3,048 women (average age 83.6±4.8; mean follow-up=5.4 years), Cox proportional hazards models examined the association of LS (actigraphy-estimated sleep duration>8h) and ISSD (insomnia [difficulty initiating or maintaining sleep and/or sleep medication use ≥3/week] and concurrent actigraphy-estimated sleep duration<6h) with mortality. Other phenotypes (insomnia with normal sleep duration [INSD; insomnia and sleep duration 6-8h]; asymptomatic short sleep [AS; no insomnia and sleep duration<6h]) were also examined. Participants with normal sleep (NS; no insomnia and sleep duration 6-8h) served as the reference group. Models were adjusted for demographics and comorbidities.

**Results:** In unadjusted models, LS was associated with increased mortality in men and women when compared with NS. In women only, LS was associated with higher mortality after adjustment for demographics and comorbidity compared with NS (HR 1.30 [1.07, 1.59]). In demographic-adjusted models and across cohorts, ISSD was significantly associated with an increased hazard of mortality compared with NS (HR 1.25 [1.10, 1.43] for men; 1.36 [1.11, 1.67] for women). This association was not significant in either cohort after adjusting for comorbidity. Persons with INSD or AS did not have increased mortality risk compared with NS.

**Conclusion:** LS and ISSD are at-risk phenotypes in older persons. Associations with mortality may be mediated by chronic diseases. Future work should examine whether sleep improvements decrease mortality in older persons.

**STATEMENT OF SIGNIFICANCE:** Prior studies have reported inconsistent results on the association between insomnia with objective short sleep duration (ISSD) and mortality in older persons. While long sleep duration (LS) has been associated with mortality, self-reported assessments may be subject to bias. Through analysis of two large cohorts of community-dwelling older men and women with objective sleep measurements and robust longitudinal data, we found ISSD and LS to be associated with all-cause mortality. After controlling for the presence of chronic conditions, however, only LS in women was significantly associated with mortality. Our findings suggest that underlying comorbidities mediate the risk of ISSD and LS in older persons and that objective measures may be needed to differentiate risk among older persons with insomnia symptoms.

## INTRODUCTION

Insomnia or poor sleep with objective short sleep duration (ISSD) has emerged as an important sleep phenotype in middle-aged persons due to its association with an elevated risk of cardiometabolic disease, cognitive impairment, and mortality among middle-aged persons.^1–4^ Recent work among older persons (>65 years of age) has indicated that ISSD is prevalent in older persons, affecting around 20% of men and 13% of women, and is cross-sectionally associated with a diverse range of health conditions, including depression, anxiety, obesity, multimorbidity, and falls.^5^ However, the prospective association of ISSD with adverse outcomes among older persons has not been evaluated.

In previous work evaluating the association of ISSD with mortality among younger and middle aged persons, results have been mixed. In the Penn State Cohort, where the average age was 50 years in men and 47 years in women, ISSD (as compared to persons without insomnia and with sleep duration ≥6 hours) was associated with increased mortality in men only.^1^ In the Sleep Heart Health Study (SHHS), where the average age was 64 years, persons with ISSD did not have an increased mortality risk when compared to persons without insomnia and sleep duration ≥6 hours.^2^ Whether ISSD increases the risk of all-cause mortality in older persons is unknown.

Notably, the aforementioned studies did not separately evaluate individuals with long sleep duration (LS), which has a well-known association with increased mortality.^6,7^ When evaluating the association of ISSD with mortality, it may be important to remove long sleepers from the reference group. Otherwise, grouping normal sleepers with long sleepers could bias estimates of the association between ISSD and mortality towards the null. In addition, the majority of studies evaluating the association of sleep duration with mortality have relied on self-reported data for sleep duration, which may be inaccurate, especially among older persons.^8^ Objective evaluation of sleep duration may be needed to accurately assess the association of sleep duration with adverse outcomes. In addition, whether sleep phenotypes are associated with increased mortality in older persons after controlling for comorbidities has not been examined.

To address these gaps, this longitudinal study investigated the association of ISSD, LS, and other sleep phenotypes with all-cause mortality in older men and women. Following prior research, we characterized ISSD based on subjective reports of insomnia from validated questionnaires and objectively-measured sleep duration.^1,2,5^ We also used objective methods to define the LS group. We hypothesized that both ISSD and LS would be associated with an increased risk of all-cause mortality in our samples of older men and women.

## METHODS

### Study Population

This is a secondary, longitudinal analysis of data from men in the Osteoporotic Fractures in Men (MrOS) Sleep Study and women in the Study of Osteoporotic Fractures (SOF).^9–11^ For both studies, inclusion criteria were age ≥65 years, no history of bilateral hip replacements and ability to ambulate without the assistance of another person.^9–11^ The study protocols were approved by the institutional review boards at all participating centers and included written informed consent.

In MrOS, 5,994 community-dwelling men were recruited from 2000-02 at six clinical centers in the United States (US).^11^ Of these, 3,135 participants were subsequently recruited from 2003-05 for the MrOS Sleep Study. In SOF, 9,704 community-dwelling Caucasian women were recruited into the original cohort from 1986-88, and 662 African-American women were purposefully recruited to increase diversity from 1997-98.^10^ Of these, 4,727 participants completed a comprehensive sleep assessment during visit 8 (2002-04). For both studies, the sleep assessment included an interview with validated sleep questionnaires and clinical measures. Shortly after the sleep study visit, participants completed wrist actigraphy through wearing a SleepWatch-O® actigraph (Ambulatory Monitoring, Inc.) on the non-dominant hand for up to seven days. Polysomnography (PSG) from one overnight unattended in-home sleep study was available in most men and in a small convenience sample in women. These data were used to identify sleep-disordered breathing (SDB). For the current study, the baseline was defined as the date of the first sleep study visit and participants were followed through February 2024. The analytical samples for both cohorts (N=3,054 in MrOS; N=3,048 in SOF) included all those who enrolled in the sleep studies with non-missing data on actigraphy, covariates and applicable items from sleep questionnaires (see *Supplemental Figure S1*).

### Demographic and Clinical Characteristics

Demographic and clinical characteristics included age, minority race (vs. White), education (less than high school vs. other), obesity (body mass index ≥30), depression (Geriatrics Depression Scale score ≥6),^12^ and history of diabetes, chronic obstructive pulmonary disease (COPD), stroke, hypertension, heart attack, or heart failure. Obesity, depression, and history of medical conditions were assessed at the time of the sleep assessment. Due to the limited availability of PSG data in women, we evaluated the presence of severe SDB in men only. SDB was established by an apnea hypopnea index (≥4% desaturation) ≥30 per hour of sleep on PSG, which has previously been shown to increase mortality risk among middle- and older-age men.^13^

### Sleep Phenotypes

We classified participants as having “insomnia or poor sleep” based on self-reported insomnia symptoms and/or sleep medication use items from the Pittsburgh Sleep Quality Index (PSQI).^14–16^ Similar to definitions previously used to evaluate ISSD,^2,5^ we classified persons as having insomnia if they reported that any of the following occurred at least three times per week: trouble getting to sleep within 30 minutes, waking up in the middle of the night or early morning, and/or taking a medication to help with sleep.

Objective overnight sleep duration was evaluated by wrist actigraphy. While PSG is the gold standard for measurement of sleep, a single night of PSG is unlikely to represent habitual sleep due to night-to-night variability and effects of sleep disruption during PSG monitoring.^17^ In addition, meta-analyses of polysomnography vs. actigraphy have shown narrow ranges in the mean difference in sleep duration between these two measures,^18^ with actigraphy measuring 13-18 more minutes on average of total sleep time in older men and women.^19,20^ Actigraphy is less susceptible to the first night effect as it is worn across multiple days and its use for estimation of sleep duration in a variety of sleep disorders is advocated by the American Academy of Sleep Medicine.^18^

Participants wore the SleepWatch-O^®^ actigraph (Ambulatory Monitoring, Inc) on the non-dominant hand. The completion rates of actigraphy were 97.4% in MrOS (mean of 5.2 [0.9] nights) and 94.8% in SOF (mean of 4.1 [0.8] nights). Actigraphy data were analyzed using Action W-2 software with Proportional Integration Mode and the University of California, San Diego scoring algorithm.^21^ Full details of these methods have been published previously.^19,21^ The software algorithm and sleep diaries were used to edit the raw data and generate variables for different sleep measures, including sleep duration. Prior analyses of actigraphy data from MrOS and SOF have shown high inter-scorer reliability.^20,22^ We defined sleep duration as normal (6 to 8 hours), short (<6 hours), or long (>8 hours) based on previously published reports examining insomnia with objective short sleep duration as well as extensive literature among older persons demonstrating adverse outcomes (mortality, impaired cognition, impaired functional capacity) with sleep durations <6 hours or >8 hours.^1,6,23–26^

We used the presence of insomnia and the actigraphy-assessed sleep duration to create five mutually exclusive categories: ISSD (insomnia and actigraphy-measured sleep duration <6 hours), LS (actigraphy-measured sleep duration >8 hours), insomnia with normal sleep duration (INSD; insomnia with actigraphy-measured sleep duration of 6-8 hours), asymptomatic short sleep (AS; no insomnia with actigraphy-measured sleep duration of <6 hours), and normal sleep (NS; no insomnia and actigraphy-measured sleep duration 6-8 hours).

### All-Cause Mortality

The outcome of this study was time-to-death, regardless of the cause (henceforth referred to as all-cause-mortality). Individuals were classified into those who died within the study period and those who were censored. For individuals who died, follow-up time was defined as the number of days between the sleep study and the date of death, based on state-registered death certificates. For individuals who were censored, follow-up time was defined as the number of days between the sleep study and the date of last contact. The last contact is defined as the latest interaction among the latest postcard contact and the most recent study visit. T mean (SD) follow-up period was 12.1 (5.6) years for men and 5.4 (2.1) years for women.

### Statistical Analysis

We analyzed men and women separately, though in parallel fashion, due to differences in demographic and clinical characteristics in the two cohorts and the unavailability of PSG in most women. ANOVA and chi-square tests were used to examine differences in demographic and clinical characteristics by sleep phenotype in each cohort. Kaplan-Meier curves with log-rank tests were used to estimate and plot the survival function associated with each of the sleep phenotypes. Subsequently, we used Cox proportional hazard models to analyze the association of the four abnormal sleep phenotypes (ISSD, LS, INSD, AS), using normal sleepers as the reference group, with all-cause mortality. For Kaplan-Meier and Cox proportional hazard models, study subjects were classified as censored (lost to follow-up, withdrawn from study or still alive at follow-up) or having experienced the event of interest (death) prior to end-of-study. Time to follow-up was defined as the number of days between date of the first sleep visit and death, for those who died during the study, or date of last contact, for those who were censored.

We examined unadjusted, demographic adjusted (adjusted for age, race, and education), and demographic and comorbidity adjusted models (additionally adjusted for obesity, depression, diabetes, COPD, stroke, hypertension, heart attack, congestive heart failure; models in men also adjusted for SDB). Plots of Schoenfeld residuals were generated for fully adjusted models to determine whether the proportionality assumption was met for all sleep phenotypes. We found no evidence that the proportional hazards assumption was violated (see *Supplemental Table S1*). Additional diagnostics included the calculation of deviance and DFBETAS statistics to detect outliers and influential observations. We also examined whether the noninformative censoring assumption was violated by comparing the hazard ratios estimated under the best- and worst-case scenarios. The best-case scenario assumes that all censored participants lived up to the end of study (16 years in MrOS; 7 years in SOF), while the worst-case scenario assumes that all censored participants died on the day lost to follow-up. All analyses were performed using SAS 9.4 (SAS Institute Inc., Cary, NC). Type I error rate of 0.05 was used to determine statistical significance.

## RESULTS

Table 1 describes the characteristics of the two cohorts overall and by sleep phenotype. The prevalence of ISSD, LS, INSD, and AS was 20.7%, 7.2%, 39.5%, and 11.2%, respectively in men, and 13.1%, 14.0%, 34.0%, and 11.9%, respectively, in women. During the study period, 72.4% of men and 33.7% of women died. Compared with normal sleepers, individuals with ISSD were less likely to be White, less educated, and had higher rates of obesity, depression, diabetes, COPD, hypertension, heart attack, and heart failure. Individuals with ISSD had a higher prevalence of comorbidities compared with INSD. Individuals with LS were older, more likely to be White, and had higher prevalence of depression and heart attack than normal sleepers. Among the LS group, 156 men (70.9%) and 264 women (61.8%) also had insomnia symptoms.

**Table 1.**
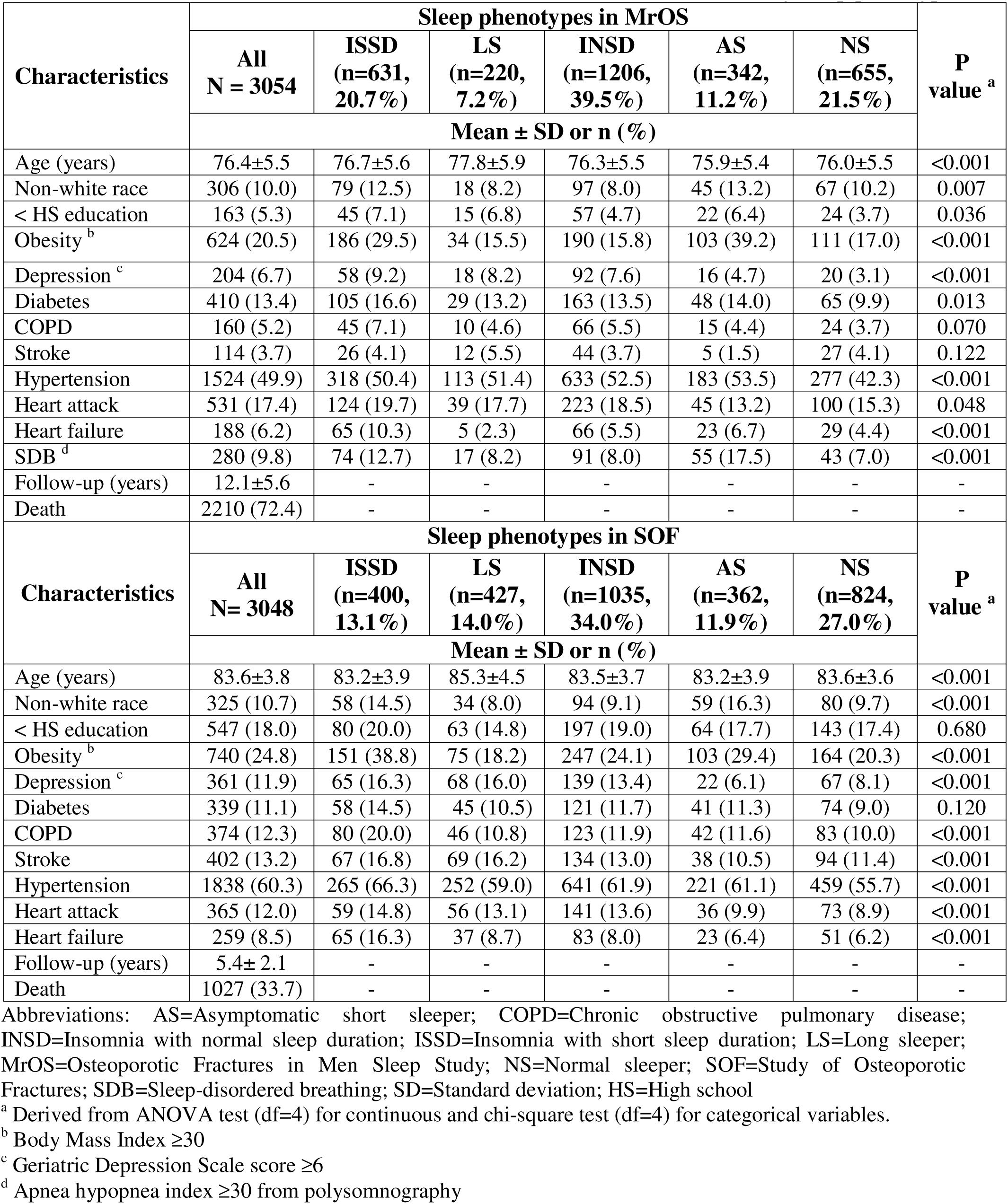
Characteristics of older men (MrOS) and women (SOF) in overall and by sleep phenotype.

Figure 1 shows Kaplan-Meier curves depicting unadjusted probabilities of survival at each time point for the five sleep phenotypes in older men and women, respectively. In both men and women, there were significant between-phenotype differences in the probability of survival over time (log-rank test, *p*s<0.05). In both men and women, LS and ISSD had the lowest probability of survival throughout the study period.

**Figure 1.**
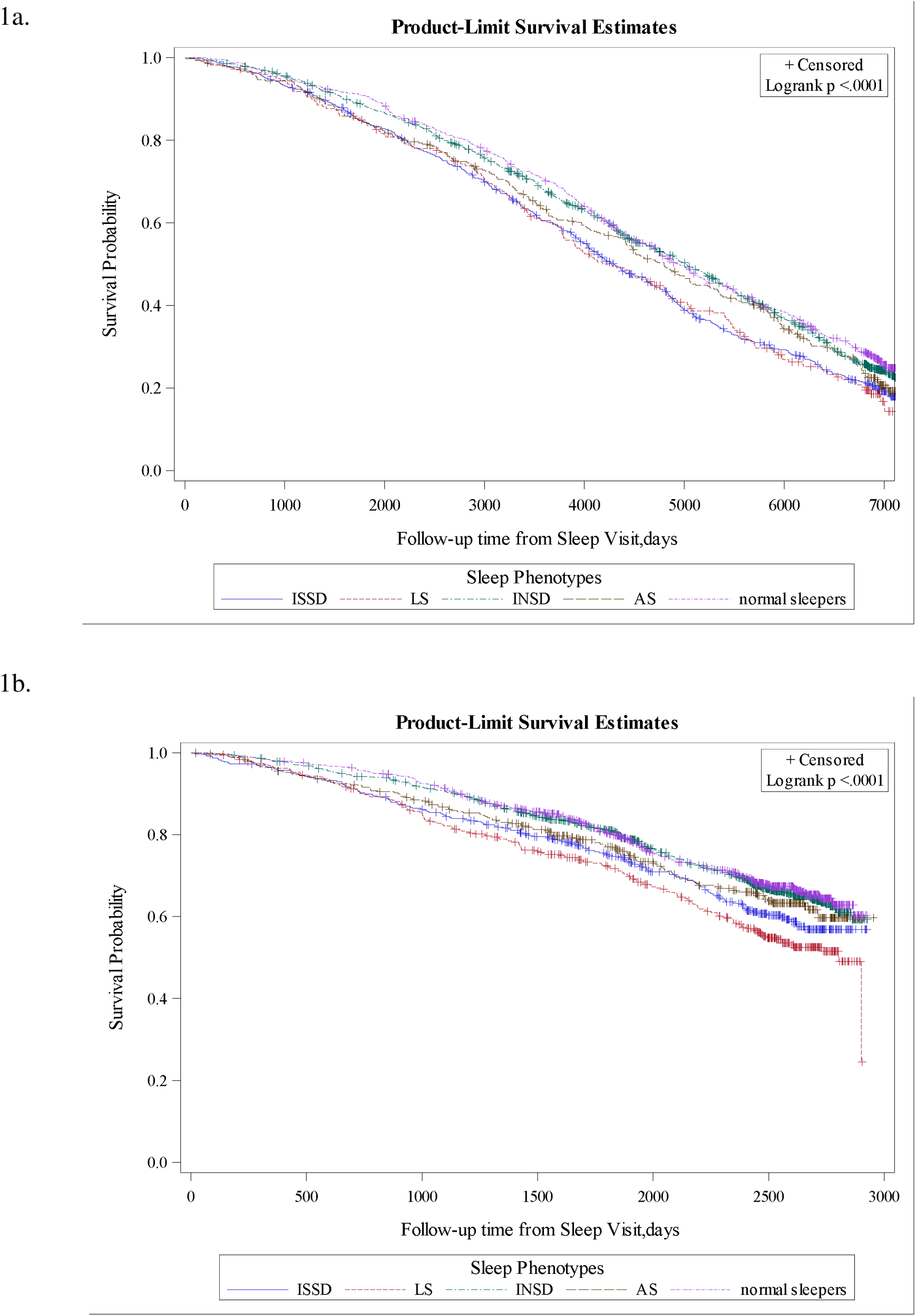
Kaplan-Meier curves of mortality according to sleep phenotype compared to normal sleepers in MrOS (Figure 1a) and SOF (Figure 1b). Figure 1a and 1b demonstrate the survival experiences of five sleep phenotypes in MrOS and SOF across the follow-up period. P-values of log-rank were <0.0001, indicating that at least one of the phenotypes has a significant difference in survival compared to the others.

Tables 2 and 3 show the risk of all-cause mortality for the four abnormal sleep phenotypes (i.e., ISSD, LS, INSD, and AS) relative to normal sleepers derived from Cox proportional hazard models. Older men with ISSD had a significantly higher risk of mortality compared to normal sleepers after controlling for age, race, and educational attainment (Model 2; HR_ISSD_, 1.25 [95% Confidence interval 1.10-1.43]). After additionally adjusting for comorbidities, the association of ISSD with mortality was attenuated and no longer significant (Model 3; HR_ISSD_, 1.12 [0.98, 1.28]). Women with ISSD and women with LS had significantly higher risks of mortality than normal sleepers after adjusting for demographic characteristics (Model 2; HR_ISSD_, 1.36 [1.11, 1.67]; HR_LS_, 1.41 [1.17, 1.71]). After additionally adjusting for comorbidities, only LS was independently associated with a higher risk of mortality than normal sleep (HR_LS_, 1.30 [1.07, 1.59]). Persons with INSD and AS did not have increased mortality risk compared with normal sleepers. Our model diagnostics did not find outliers or influential observations in analyses of either cohort. We also did not find the assumption of noninformative censoring to be violated from our analysis of the best- and worst-case scenarios (see *Supplemental Table S2*).

**Table 2.** Association between abnormal sleep phenotypes and all-cause mortality among older men, MrOS.

| Characteristics <sup>a</sup> | Older men (MrOS)<br>N=3054 |  |  |
| --- | --- | --- | --- |
|  | Model 1 <sup>b</sup> | Model 2 <sup>c</sup> | Model 3 <sup>d</sup> |
|  | Hazard Ratios (95% CI) |  |  |
| ISSD | **1.29 (1.14, 1.47)** | **1.25 (1.10, 1.43)** | 1.12 (0.98, 1.28) |
| AS | 1.15 (0.99, 1.34) | **1.24 (1.06, 1.44)** | 1.13 (0.96, 1.33) |
| INSD | 1.05 (0.94, 1.18) | 1.01 (0.90, 1.13) | 0.97 (0.86, 1.09) |
| LS | **1.33 (1.11, 1.58)** | 1.13 (0.95, 1.35) | 1.03 (0.86, 1.24) |

Hazard Ratios of statistical significance were bolded.
Abbreviations: AS=Asymptomatic short sleepers; INSD=Insomnia with normal sleep duration; ISSD=Insomnia with short sleep duration; LS=Long sleepers; MrOS=Osteoporotic Fractures in Men Sleep Study; COPD=Chronic obstructive pulmonary disease; SDB=Sleep-disordered breathing
<sup>a</sup> See footnotes to Table 1 regarding definitions for characteristics.
<sup>b</sup> From unadjusted proportional hazards models
<sup>c</sup> From proportional hazards models adjusting for age, race, education
<sup>d</sup> From proportional hazards models adjusting for age, race, education, obesity, depression, diabetes, COPD, stroke, hypertension, heart attack, heart failure, and SDB.

**Table 3.** Association between sleep phenotypes and all-cause mortality among older women.

| Characteristics <sup>a</sup> | Older women (SOF)<br>N=3048 |  |  |
| --- | --- | --- | --- |
|  | Model 1 <sup>b</sup> | Model 2 <sup>c</sup> | Model 3 <sup>d</sup> |
|  | Hazard Ratios (95% CI) |  |  |
| ISSD | **1.31 (1.07, 1.61)** | **1.36 (1.11, 1.67)** | 1.14 (0.92, 1.42) |
| AS | 1.17 (0.94, 1.46) | 1.25 (1.00, 1.56) | 1.25 (1.00, 1.58) |
| INSD | 1.04 (0.88, 1.22) | 1.04 (0.89, 1.23) | 0.97 (0.82, 1.15) |
| LS | **1.56 (1.29, 1.88)** | **1.41 (1.17, 1.71)** | **1.30 (1.07, 1.59)** |

Hazard Ratios of statistical significance were bolded.
Abbreviations: AS=Asymptomatic short sleepers; INSD=Insomnia with normal sleep duration; ISSD=Insomnia with short sleep duration; LS=Long sleepers; MrOS=Osteoporotic Fractures in Men Sleep Study; COPD=Chronic obstructive pulmonary disease
<sup>a</sup> See footnotes to Table 1 regarding definitions for characteristics.
<sup>b</sup> From unadjusted proportional hazards models
<sup>c</sup> From proportional hazards models adjusting for age, race, education
<sup>d</sup> From proportional hazards models adjusting for age, race, education, obesity, depression, diabetes, COPD, stroke, hypertension, heart attack, and heart failure.
AS= Asymptomatic short sleepers
COPD= Chronic obstructive pulmonary disease

## DISCUSSION

In this large sample of community-dwelling older men and women, persons with LS had an increased risk of all-cause mortality compared with normal sleep in unadjusted models, and this relationship remained significant in women only after further adjusting for the presence of chronic comorbidities. Individuals with ISSD had an increased risk of all-cause mortality after controlling for age, race, and educational attainment. These associations were attenuated and no longer significant after adjusting for the presence of chronic conditions (i.e., depression, cardiometabolic conditions, and SDB [in men only]), suggesting that these associations were explained by greater comorbidity burden among individuals with ISSD.

In previous studies, findings for the association of ISSD with all-cause mortality have been mixed.^4,5^ In the Penn State cohort, ISSD (defined by reporting insomnia symptoms lasting ≥1 year and sleep duration <6h by PSG) was significantly associated with all-cause mortality in men but not women in analyses adjusted for demographics, BMI, hypertension, diabetes, depression, smoking, alcohol use, and SDB (apnea hypopnea index ≥5 events/hr).^5^ In SHHS, ISSD (where insomnia was defined as in the current study and short sleep duration as <6h by PSG) was not significantly associated with all-cause mortality in propensity score-adjusted analyses that included men and women.^4^ Age differences may partially explain the observed differences in the Penn State cohort, where the average was 50 years in men, versus average ages of 64 and >75 years in SHHS and the current study, respectively. Middle-aged men with ISSD may have hyperactivation of the hypothalamic-pituitary-adrenal axis, leading to exacerbations of health (e.g., diabetes, hypertension) that increase mortality risk. Middle-aged women may be less susceptible to hyperactivation of the HPA axis, though the authors in the Penn State study acknowledge that sex differences may have resulted from a smaller sample size and shorter follow-up period in women. It is also possible that the definition for insomnia in the Penn State cohort selected for a more severe and chronic form of insomnia as compared with SHHS and the current study. Our results align with those reported in SHHS, suggesting that ISSD among older persons may be an at-risk phenotype but is not independently associated with all-cause mortality. Rather, the association of ISSD with mortality may be explained by a higher burden of chronic diseases among those with ISSD. Hence, future research should determine if identifying and intervening on ISSD is an efficacious strategy for reducing the burden of morbidity and mortality in older persons. It may also be important to examine whether short sleep due to fragmentation or reduced sleep opportunity among persons with ISSD is differentially associated with adverse outcomes.

Another interesting finding of the current study, supported by results in the Penn State cohort and SHHS, was that objective short sleep duration, in the absence of insomnia symptoms, was not associated with increased mortality. This finding contrasts with a large body of research suggesting a U-shaped association between sleep duration and mortality. However, most studies supporting a U-shaped association relied on self-reported measures to assess sleep duration. Many demographic, psychosocial, and health factors are associated with under- or over-reporting of sleep compared with objective measures, introducing systemic bias and inappropriate causal inference (i.e., it may be that factors associated with perceived short sleep confound the association between short sleep duration and mortality).^27^ Thus, our work and the work of others suggests that objective measures may be needed to accurately assess short sleep duration as a risk factor for adverse outcomes.^1,2,27^ Our results are consistent with recent work by Mendoza-Fernandez and others that propose that insomnia is a highly heterogeneous condition and best characterized by describing subtypes that utilize both symptoms and objective data.^28^ Similar to prior findings, we did not identify an increased risk of mortality for insomnia with objectively measured normal sleep duration.^1,2^ Whether the sleep phenotypes studied here are associated with adverse outcomes other than mortality should be investigated in future work.

Our findings extend previous work by examining objectively-defined LS as a high-risk phenotype, offering a clearer picture of the association of long sleep duration with mortality among older persons. Although many studies have identified a link between long sleep and increased mortality risk, this connection is often more pronounced in self-reported data, which is subject to bias in reporting.^27^ Perceived long sleep duration may be a less reliable indicator of actual sleep and, consequently, its association with mortality may be more questionable. Our results demonstrate an increased mortality risk among older men and women with objective increased sleep duration (>8 hours), with a robust association in older women, but not men, after controlling for demographics and cardiometabolic conditions. The robust association in women may have resulted from the older age and higher prevalence of LS in women. Whether long sleep is a causal factor or simply a marker of underlying illness remains to be fully elucidated. However, it seems plausible, especially among persons of advanced age, that underlying conditions (e.g., chronic inflammatory conditions, sleep-fragmenting disorders, neurodegenerative diseases) simultaneously drive both extended sleep and increased mortality. The presence of such underlying conditions may also explain the sex differences demonstrated here, since women are more likely to experience autoimmune and chronic inflammatory conditions and are more likely to develop Alzheimer’s disease.

The current work and that of others suggests that relying solely on self-reported sleep duration can lead to misclassification of sleep disorder severity. This work has several implications. First, when encountering persons with insomnia, objective evaluation of sleep duration may allow personalized treatments and avoid over-medicalization. Judicious use of wearable devices may complement symptom reports to identify both informative subtypes and personalize interventions (i.e., using sleep duration data) to create appropriate treatment plans. For persons with long sleep duration, our results confirm what has been suggested by self-reported studies and indicate that long sleep duration, whether perceived or objectively identified, should trigger practitioners to identify and manage underlying conditions.

Our study has several strengths. First, we focused on community-dwelling older persons with a representative sample of those in middle- and oldest old,^29^ an understudied population with a high prevalence of sleep disturbances. Second, we used actigraphy to assess objective sleep duration. Actigraphy is more accurate in assessing sleep duration than self-report, is less susceptible to the first-night effect, and is more easily deployed as an objective measure of sleep than PSG. Third, we used survival analysis accompanied by rigorous testing of the proportionality assumption, model diagnostics, and sensitivity analysis. Fourth, we evaluated a broad range of covariates and compared risk estimates across multiple adjusted models, which enabled us to explore possible mediating pathways. Fifth, we separately evaluated long sleepers, which may allow more accurate estimates of mortality risk associated with abnormal sleep phenotypes.

Nevertheless, we acknowledge several limitations. First, we were unable to account for SDB in older women, as the majority did not undergo PSG. While accounting for this condition may attenuate the association of long sleep with mortality, previous work suggests that SDB is not independently associated with increased all-cause mortality in women.^13^ Second, our definition of “insomnia or poor sleep” is not synonymous with a diagnosis of insomnia disorder, which considers chronicity and daytime impairments related to insomnia symptoms. Nevertheless, our definition matches that used previously in SHHS, allowing close comparisons between the two studies. It may be that using a diagnosis of insomnia disorder, rather than insomnia symptoms, to define ISSD would be associated with a higher mortality risk. Third, sleep phenotypes were defined cross-sectionally and do not account for changes in an individual’s sleep quality over time. Fourth, while actigraphy allows an estimate of sleep duration from multiple nights of measurement, it overestimates sleep duration overall and has worse performance among persons with poor sleep.^19,20,30^ Thus, there may be misclassification of sleep duration in the normal sleep duration groups, which would bias towards null results. Nonetheless, PSG measures of sleep duration also have limitations.^17^ Devices that are better at measuring habitual sleep in the home setting, especially among poor sleepers, are needed to overcome the limitations of PSG and ACT. Finally, our analyses do not account for daytime sleep, which may buffer against mortality risk in persons with ISSD.

In conclusion, in a large cohort of community-dwelling older men and women, we found ISSD and LS to be at-risk phenotypes associated with all-cause mortality in unadjusted analyses, while LS was independently associated with mortality in women, but not men, after controlling for chronic conditions. Our findings suggest that objective measurements may be needed to accurately assess sleep duration as a risk factor for adverse outcomes and guide clinical management of sleep disturbances, particularly in cases of perceived short sleep duration. Future studies are needed to investigate pathways underlying the association of ISSD and LS with mortality and to determine whether sleep interventions might reduce morbidity and mortality among older persons.

## Data Availability

All data produced are available online at https://mrosonline.ucsf.edu and https://sofonline.ucsf.edu.

## Acknowledgements

The Osteoporotic Fractures in Men (MrOS) Study is supported by National Institutes of Health funding. The following institutes provide support: the National Institute on Aging (NIA), the National Institute of Arthritis and Musculoskeletal and Skin Diseases (NIAMS), and the National Center for Advancing Translational Sciences (NCATS) under the following grant numbers: U01 AG027810, U01 AG042124, U01 AG042139, U01 AG042140, U01 AG042143, U01 AG042145, U01 AG042168, U01 AR066160, R01 AG066671, and UL1 TR002369.

The Study of Osteoporotic Fractures (SOF) is supported by National Institutes of Health funding. The National Institute on Aging (NIA) provides support under the following grant numbers: R01 AG005407, R01 AR35582, R01 AR35583, R01 AR35584, R01 AG005394, R01 AG027574, R01 AG027576, and R01 AG026720.

## Data Sharing Statement

The data supporting this study will be made available upon reasonable request by contacting the corresponding author.

## Abbreviations

AS: Asymptomatic short sleepers
CI: Confidence interval
COPD: Chronic obstructive pulmonary disease
HR: Hazard ratio
INSD: Insomnia with normal sleep duration
ISSD: Insomnia with short sleep duration
LS: Long sleepers
MrOS: Osteoporotic Fractures in Men Sleep Study
PSG: polysomnograph
SDB: Sleep-disordered breathing
SD: Standard deviation
SHHS: Sleep Heart Health Study
SOF: Study of Osteoporotic Fractures

