## Supplemental materials for "Association of Objective Long Sleep Duration and Insomnia with Objective Short Sleep Duration Phenotypes with Mortality in Older Persons"

Figure S1. Analytic sample derived from MrOS and SOF

(A)

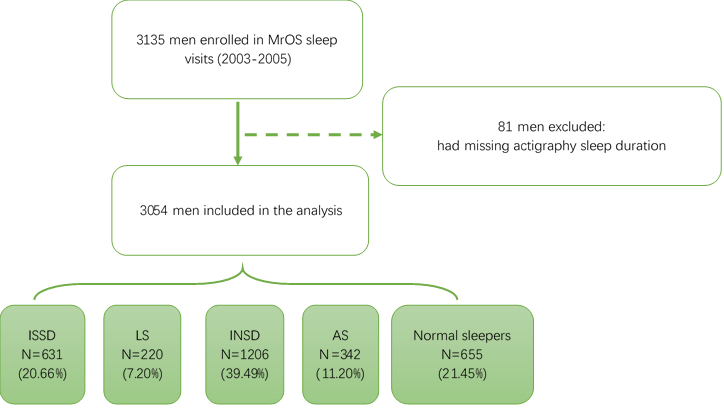

(B)

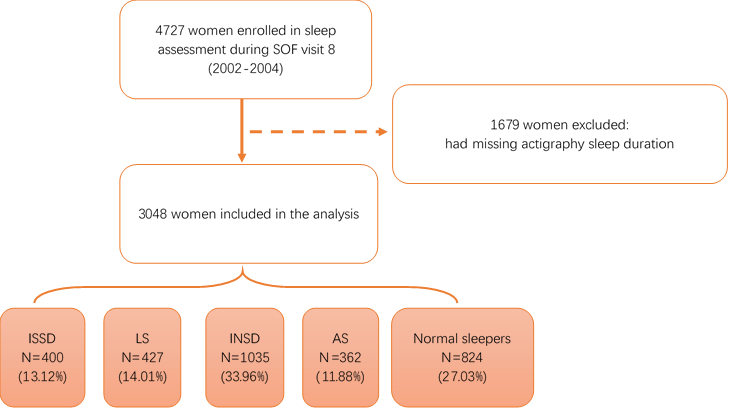

**Table S1.** Analysis of Maximum Likelihood Estimates of proportional hazard model in older men and women

| **Characteristics** | **Men (MrOS)**  **N=3054** | | **Women (SOF)**  **N=3048** | |
| --- | --- | --- | --- | --- |
|  | **Hazard Ratios** | **Pr > ChiSq** | **Hazard Ratios** | **Pr > ChiSq** |
| ISSD | 1.52 | 0.005 | 1.87 | 0.007 |
| AS | 1.26 | 0.200 | 1.75 | 0.024 |
| INSD | 1.01 | 0.920 | 1.05 | 0.804 |
| LS | 1.44 | 0.077 | 1.95 | 0.003 |
| ISSD*time | 1.00 | 0.217 | 1.00 | 0.092 |
| AS*time | 1.00 | 0.579 | 1.00 | 0.074 |
| INSD*time | 1.00 | 0.765 | 1.00 | 0.938 |
| LS*time | 1.00 | 0.661 | 1.00 | 0.266 |

Abbreviations: AS=Asymptomatic short sleepers; INSD=Insomnia with normal sleep duration; ISSD=Insomnia with short sleep duration; LS=Long sleepers; MrOS=Osteoporotic Fractures in Men Sleep Study; SOF=Study of Osteoporotic Fractures.

**Table S2.** Sensitivity analysis using best- and worst-case scenarios

| **Sleep Phenotypes ^a^** | **MrOS**  **N=3054** | | **SOF**  **N=3048** | |
| --- | --- | --- | --- | --- |
|  | **Best-case ^b^** | **Worst-case ^c^** | **Best-case ^b^** | **Worst-case ^c^** |
|  | **Hazard Ratios (95% CI)** | | | |
| ISSD | 1.08 (0.94, 1.24) | 1.14 (1.00, 1.29) | 1.13 (0.91, 1.40) | 1.13 (0.98, 1.32) |
| AS | 1.09 (0.93, 1.28) | 1.15 (0.98, 1.34) | 1.17 (0.93, 1.47) | **1.19 (1.02, 1.39)** |
| INSD | 0.93 (0.83, 1.04) | 1.00 (0.90, 1.12) | 0.97 (0.82, 1.15) | 0.97 (0.87, 1.09) |
| LS | 0.98 (0.81, 1.17) | 1.07 (0.90, 1.28) | **1.29 (1.05, 1.57)** | 1.12 (0.97, 1.29) |

Hazard Ratios of statistical significance were bolded.

Abbreviations: AS=Asymptomatic short sleepers; INSD=Insomnia with normal sleep duration; ISSD=Insomnia with short sleep duration; LS=Long sleepers; MrOS=Osteoporotic Fractures in Men Sleep Study; SOF=Study of Osteoporotic Fractures.

a. See footnotes to Table 1 regarding definitions.
b. Best-case scenario assumes all censoring participants lived to end of follow-up. From proportional hazards models adjusting for age, race, education, obesity, depression, diabetes, COPD, stroke, hypertension, heart attack, heart failure, and SDB (in men only).
c. Worst-case scenario in MrOS and SOF assumes all censoring participants died at time of censoring. From proportional hazards models adjusting for age, race, education, obesity, depression, diabetes, COPD, stroke, hypertension, heart attack, heart failure, and SDB (in men only).
